# Radiographically identified vertebral fractures in haemochromatosis-associated *HFE* C282Y homozygotes in the UK Biobank

**DOI:** 10.64898/2026.08.19.26360796

**Authors:** Lucy R. Banfield, Luke C. Pilling, David Melzer, Jeremy D. Shearman, Karen M. Knapp, Janice L. Atkins

**Affiliations:** The Department of Health and Care Professions, Faculty of Health and Life Sciences, University of Exeter, St Luke’s Campus, Exeter, UK; Department of Clinical and Biomedical Sciences, Faculty of Health and Life Sciences, University of Exeter, St Luke’s Campus, Exeter, UK; Emeritus Professor (retired), University of Exeter, St Luke’s Campus, Exeter, UK; Department of Gastroenterology, South Warwickshire University NHS Foundation Trust, Warwick, UK

**Keywords:** Vertebral fracture, C282Y homozygote, Haemochromatosis, Iron overload

## Abstract

**Purpose:** Haemochromatosis due to *HFE*-C282Y homozygosity can lead to excess iron absorption and is typically associated with liver malignancy, plus widespread arthritis. Recent evidence suggests that limb fractures are more common, but little is known about vertebral effects. This study investigated the association of vertebral compression fractures, assessed with intelligent dual-energy X-ray absorptiometry (iDXA), and *HFE* genotype in a large community cohort.

**Methods:** UK Biobank data from 227 European genetic ancestry C282Y homozygotes (mean 64.6 years) and 234 age, sex, and BMI-matched controls without common *HFE* haemochromatosis variants were included. Lateral vertebral assessment scans (iDXA, GE-Lunar) were acquired at imaging reassessment (2014-2020) and reviewed, blind to genotype, for radiological evidence of vertebral fracture. Matched logistic regression models assessed associations between C282Y homozygosity and vertebral fractures.

**Results:** 78 vertebral fractures (16.9%) were identified within 461 participants. Male C282Y homozygotes had increased odds of vertebral fracture (n=22/89, 24.7%) compared to participants without *HFE* alleles (n=9/90, 10.0%); Odds Ratio [OR]: 2.95, 95%CI: 1.28–6.85, p=0.01. The association persisted after excluding individuals with a diagnosis of haemochromatosis (OR: 3.37, 95% CI: 1.41–8.10, p=0.007). No excess fracture risk was observed in female C282Y homozygotes (n=23/138, 16.7%) vs those without *HFE* alleles (n=24/144, 16.7%); OR: 0.99, 95%CI: 0.53-1.87, p=1.00.

**Conclusion:** In this community-based imaging study, male *HFE* C282Y homozygotes had a markedly higher likelihood of vertebral fractures than those without *HFE* variants. These findings support further evaluation of vertebral fracture assessment in C282Y homozygous men to ensure prompt treatment to prevent future fracture if appropriate.

## Introduction

Haemochromatosis is an autosomal recessive disorder with the main risk variant being *HFE* C282Y homozygosity, which occurs in approximately 0.6% (around 1 in 150) of individuals of Northern European ancestry (1,2). It results in inappropriately low hepcidin levels and excessive dietary iron absorption (3). The consequent cellular iron overload leads to severe complications such as liver cirrhosis, hepatocellular carcinoma, widespread arthritis, and dementia (1,4–6). Male C282Y homozygotes typically accumulate iron earlier than females and experience greater morbidity and mortality (2,6). A second *HFE* variant (H63D) has been historically associated with the diagnosis of haemochromatosis in a small number of individuals but is associated with much weaker effects on cellular and systemic iron metabolism and related clinical outcomes than C282Y homozygosity (6,7).

Iron overload has the potential to disrupt bone remodelling and reduce bone mineral density (BMD) through several mechanisms, including the stimulation of osteoclasts and the inhibition of osteoblasts (8). Consistent with this, haemochromatosis has been associated with osteopenia, osteoporosis (9–12) and fractures (13–17). We previously reported excess musculoskeletal morbidity in male C282Y homozygotes in UK Biobank, including increased risks of osteoarthritis, joint replacement, osteoporosis and femoral fracture (6,14).

Altered bone remodelling may increase susceptibility to vertebral fragility before overt clinical manifestations become apparent. This is important because vertebral fractures strongly predict future skeletal injury, independent of age and BMD (18,19), forming part of the recognised “vertebral fracture cascade” (20). Vertebral fractures are associated with substantial morbidity, disability and mortality (21,22), yet an estimated 65–80% remain clinically unrecognised (23). Haemochromatosis has increasingly been linked to skeletal fragility, including reports of multiple vertebral fractures occurring in the absence of classic osteoporosis risk factors (13,15,24). A small study reported a non-significant association between haemochromatosis and vertebral fracture (OR 1.70, 95% CI 0.80–3.80) but relied on self-reported fractures in 306 clinically diagnosed patients and may therefore have underestimated fracture prevalence (25). However, the burden of radiographically detected vertebral fractures in genetically defined C282Y homozygotes remains uncertain. We therefore examined the association between *HFE* genotype and vertebral fractures identified using iDXA vertebral fracture assessment in UK Biobank participants.

## Methods

### Study sample

UK Biobank (UKB) is a cohort study of ∼500,000 UK adults aged 37-73 years at baseline assessment (2006 to 2010) (26). Data include baseline characteristics, biomarkers, genetics, and linked medical records. UKB data are available to bona fide researchers upon application. The Northwest Multi-Centre Research Ethics Committee approved the collection and use of UKB data (Research Ethics Committee reference 11/NW/0382). Participants provided informed consent to the use of their data, health records, and biological materials for health-related research. Access to UKB was granted under application number 14631.

### Genotype data

Genotype information on *HFE* C282Y (rs1800562 A allele) and *HFE* H63D (rs1799945 G allele) were from whole-exome sequencing data (methods by Regeneron(27)). Patient consent did not include providing patients with feedback on genotypes. We included 451,270 participants genetically similar to the 1,000 Genomes Project European Ancestry superpopulation (“EUR-like”)(28) of which 2,902 (0.64%) were C282Y homozygotes.

### IDXA Data

During follow-up, a subset of UK Biobank participants (n=41,594/451,270 at the time of analysis) attended a follow-up visit for iDXA imaging (2014 to 2020, aged 45 to 82 years). As part of a suite of imaging examinations, iDXA high-resolution posteroanterior imaging of the lumbar spine, and a lateral vertebral fracture assessment image of the thoracic and lumbar spine were performed using a GE Lunar (Madison, WI)(29).

Of the UK Biobank participants with iDXA imaging data, all available *HFE* C282Y homozygotes were identified and matched 1:1 with controls without C282Y or H63D alleles, based on age, sex, and body mass index (BMI) category. The initial matched sample comprised 472 participants. Following image review, a small number of participants were excluded due to missing lateral spine images or images of insufficient quality to permit reliable vertebral fracture assessment. This resulted in a final analytic sample of 461 participants for the vertebral fracture analysis (Figure 1). Femoral neck BMD data were not available for all participants at the relevant assessment visit, thereby reducing the sample size for BMD analyses (n=426). Consequently, participant numbers vary slightly across analyses depending on data availability.

**Figure 1.**
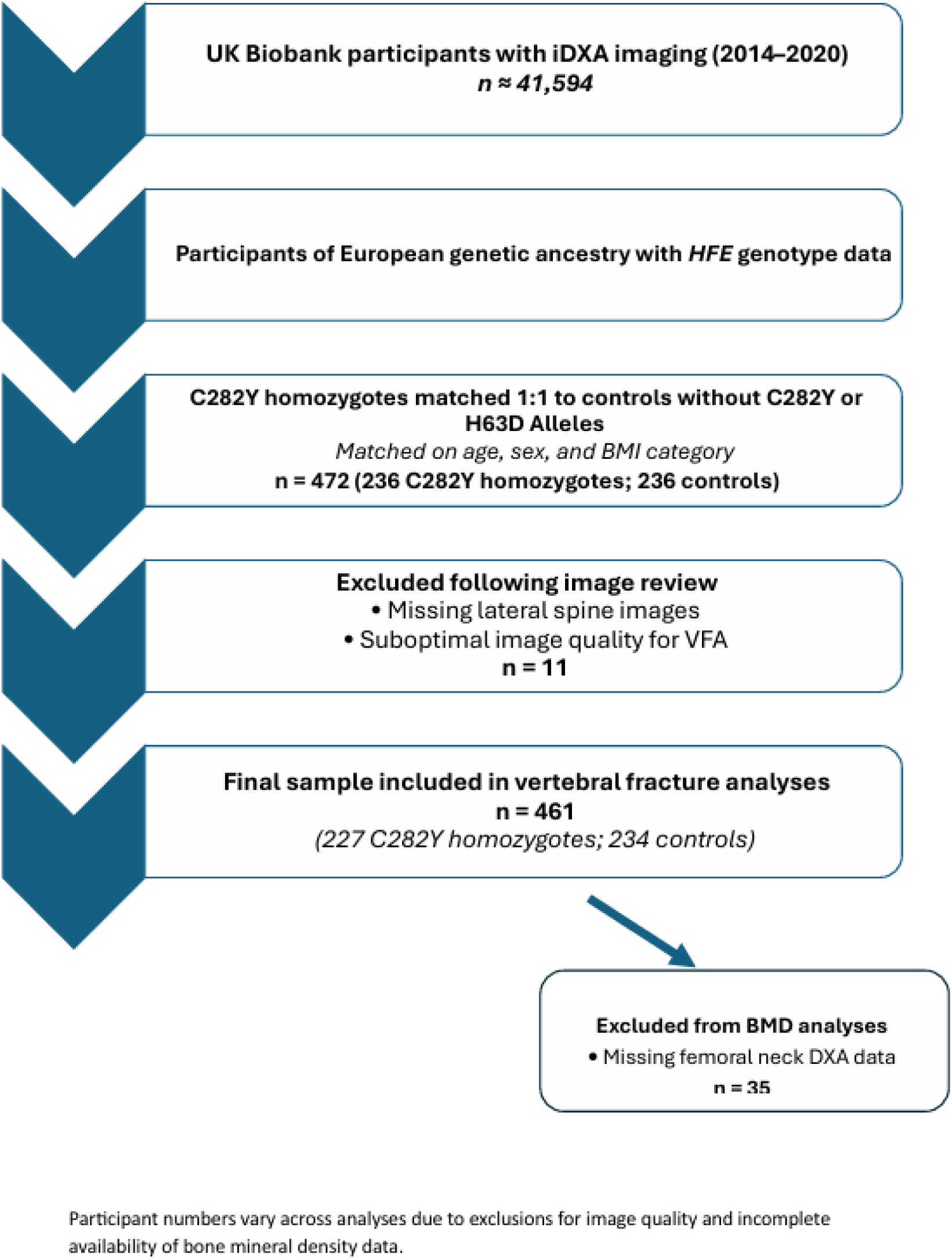
Flowchart of included participants.

Lateral vertebral fracture assessment (VFA) images acquired using iDXA were reviewed for radiological evidence of vertebral fracture by an experienced reporting radiographer (LRB), blind to participant genotype and clinical data, to minimise observer bias. For each participant, the presence, number and vertebral level of fractures were recorded. Formal Genant grading was not applied; instead, vertebral fractures were identified morphometrically and recorded as a binary outcome (fracture present or absent) to maximise reliability and minimise misclassification, given the known limitations of iDXA-based VFA in detecting mild abnormalities (30). Images deemed insufficient for reliable fracture assessment due to positioning, artefact or limited visualisation were excluded prior to analysis.

### Additional variables

Participants were asked to self-report at baseline if they had experienced back pain for 3 months or more. The prevalence of diagnosed vertebral fracture, haemochromatosis, osteoarthritis and osteoporosis was based on self-report and diagnoses from hospital inpatient data from 1996 to the date of the iDXA visit, as previously coded (Table 1) (14). BMD was assessed using a T-score derived from femoral neck iDXA scanning. Femoral neck T-scores were considered more representative of BMD, as lumbar spine data can be affected by degenerative changes and require careful correlation with imaging (31).

**Table 1:** Disease coding using ICD-10: International Classification of Diseases, Tenth Revision (World Health Organisation, 2019 version).

| **Disease** | **ICD-10 Codes** |
| --- | --- |
| Haemochromatosis | E83.1 |
| Osteoarthritis | M15.0; M15.1; M15.2; M15.9; M16.0; M16.1; M17.0; M17.1; M18.0; M18.1; M19.0 |
| Osteoporosis | M80; M80.5; M80.8; M81; M81.5; M81.8; M81.9 |
| Vertebral Fracture | M48.4; S22.0; S22.1; S32.0; T08 |

### Statistical analysis

*HFE* C282Y homozygotes were matched to controls (without C282Y or H63D alleles) for sex, age and BMI category at the time of DXA - underweight (<18.5kg/m^2^), normal (18.5-24.9kg/m^2^), overweight (25-29.9kg/m^2^) or obese (≥30kg/m^2^). We used random-effects logistic regression models, stratified by sex, to estimate associations between genotype and the likelihood of vertebral fracture using the STATA function ‘xtlogit’, specifying pairwise matching as the random intercept to account for the within-pair correlation. This approach was chosen because our study used a matched case-control design. The xtlogit model is particularly suitable for analysing matched-pair data with binary outcomes, effectively accounting for within-subject correlation and unobserved heterogeneity through random-effects models (32). We examined the prevalence of diagnosed vertebral fracture, haemochromatosis, osteoarthritis and osteoporosis, and self-reported back pain within participants by genotype group. We also performed an additional sensitivity analysis, excluding individuals diagnosed with haemochromatosis at the time of iDXA imaging (n=35). The majority of analyses were carried out in STATA 18.0 using a matched analysis. Additional analyses were performed using R within the UK Biobank Research Analysis Platform (RAP), a secure cloud-based Trusted Research Environment providing access to UK Biobank data.

## Results

### Characteristics of participants

Analyses included 461 participants of European descent aged 48 to 80 years at the time of iDXA scanning (227 C282Y homozygotes and 234 controls). The mean age was 64.6 years (SD ±7.6) and women comprised 61.2% of the sample (n=282) (Table 2).

**Table 2.** UK Biobank participant characteristics.

| Group |  | *HFE* genotype | N | Mean age, years (SD) | Haemochromatosis diagnosis at time of DXA (%) | OA at time of DXA (%) | OP at time of DXA (%) | Back pain for 3+ months (%) |
| --- | --- | --- | --- | --- | --- | --- | --- | --- |
| Males | No vertebral fracture | Non-carriers* | 81 | 64.27 (7.80) | 0 | 7 (8.6) | <5 (<6.2%) | 9 (11.1) |
|  |  | C282Y homozygotes | 67 | 64.85 (7.80) | 19 (28.4) | <5 (<7.5) | 0 | 10 (14.9) |
|  | Vertebral fracture | Non-carriers* | 9 | 67.67 (8.19) | 0 | <5 (<55.6) | 0 | <5 (<55.6) |
|  |  | C282Y homozygotes | 22 | 64.41 (6.75) | <5 (<22.7) | 0 | 0 | <5 (<22.7) |
| Females | No vertebral fracture | Non-carriers* | 120 | 64.19 (7.51) | 0 | 13 (10.8) | <5 (<4.2%) | 15 (12.5) |
|  |  | C282Y homozygotes | 115 | 64.38 (7.56) | 10 (8.7) | 10 (8.7) | 0 | 18 (15.7) |
|  | Vertebral fracture | Non-carriers* | 24 | 67.08 (6.01) | 0 | <5 (<20.8) | <5 (<20.8%) | <5 (<20.8) |
|  |  | C282Y homozygotes | 23 | 67.35 (7.97) | <5 (<21.7) | <5 (<21.7) | 0 | <5 (<21.7) |

Abbreviations: OA, Osteoarthritis. OP, Osteoporosis.
The ‘<’ symbol is used in accordance with UK Biobank’s data disclosure policy, which prohibits reporting exact cell counts smaller than 5 to protect participant confidentiality.
\* Non-carriers = no *HFE* C282Y or H63D alleles.

### Vertebral fracture

Overall, vertebral fractures were identified in 78 (16.9%) participants, of whom 45 were C282Y homozygotes. In males, vertebral fractures were present in 24.7% of C282Y homozygotes (22/89) and 10.0% without *HFE* variants (9/90). In females, vertebral fractures were present in 16.7% of C282Y homozygotes (23/138) and 16.7% without *HFE* variants (24/144). (See Figure 2 for examples of vertebral fractures identified in the imaging data). The mean age of those with vertebral fractures was marginally lower among male homozygotes (64.4 years) than among matched controls without *HFE* alleles (67.7 years). However, there was no appreciable difference in mean age between female C282Y homozygotes and their matched controls (67.4 and 67.1 years, respectively) in those with vertebral fractures.

**Figure 2.**
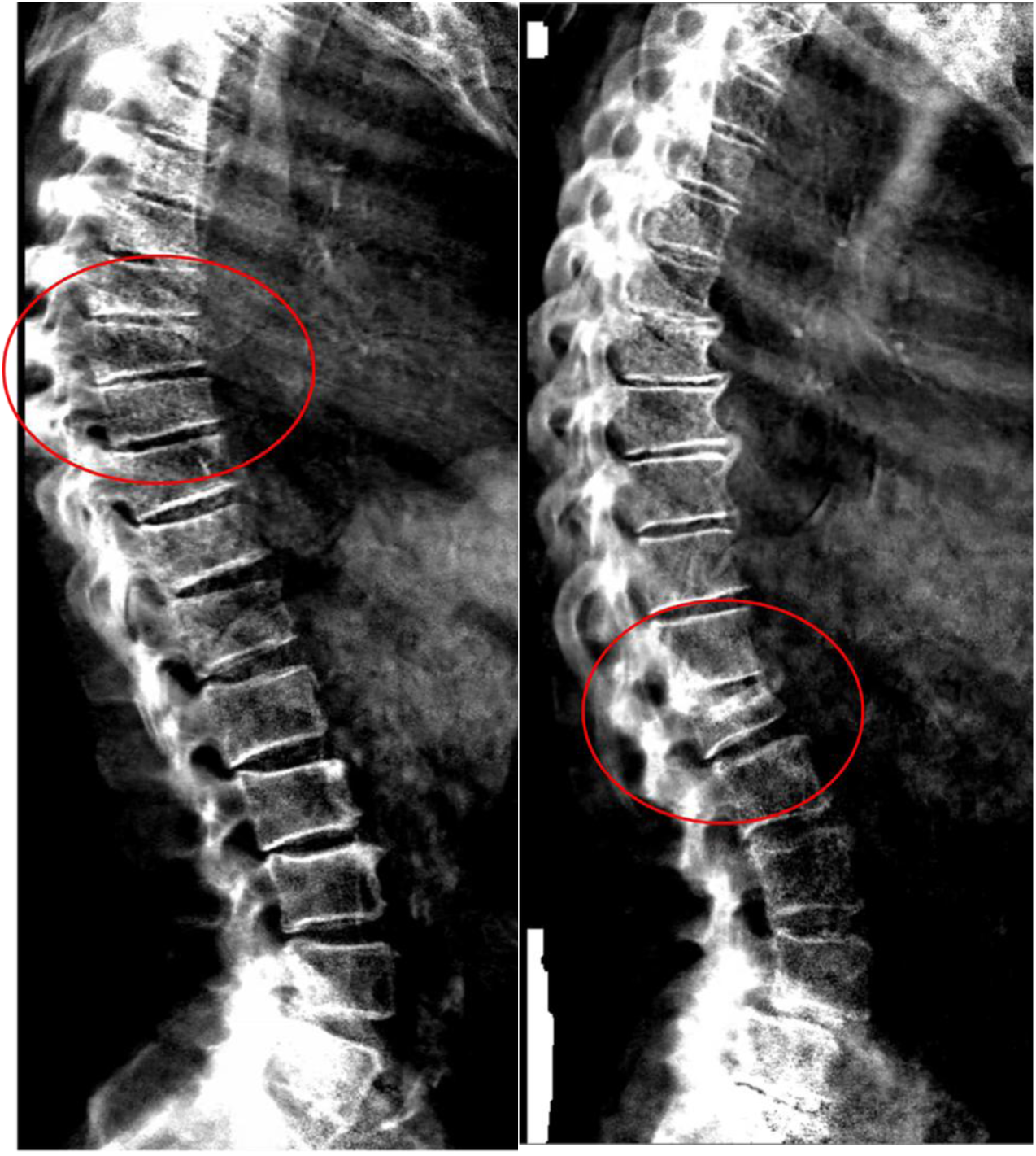
iDXA images of previously undiagnosed vertebral fractures in two randomly selected male C282Y homozygotes within UK Biobank. Reproduced by kind permission of UK Biobank©

Among participants with an identified vertebral fracture, 26 (33.3%) had fractures at more than one vertebral level. By contrast, vertebral fractures were infrequently diagnosed in hospital episode data for this cohort (n<5), highlighting a substantial gap between fractures detected on imaging and those captured in routine clinical records.

### Regression Analysis

Sex-stratified matched regression analysis demonstrated increased odds of vertebral fracture in male C282Y homozygotes compared with age- and BMI-matched controls without *HFE* C282Y or H63D alleles (OR: 2.95, 95% CI: 1.28–6.85, p=0.01). In contrast, no increase in vertebral fracture odds was observed among female C282Y homozygotes compared with matched controls (OR: 0.99, 95% CI: 0.53–1.87, p=1.00) (Table 3).

**Table 3.** Odds of vertebral fracture by C282Y genotype status.

| **Group** | **HFE genotype** | **N** | **Vertebral Fracture, n (%)** | **OR** | **95% CIs** | **p-value** |
| --- | --- | --- | --- | --- | --- | --- |
| **Males** | **Non-carriers\*** | 90 | 9 (10.0) | 1.00 |  |  |
|  | **C282Y homozygotes** | 89 | 22 (24.7) | 2.95 | 1.28–6.85 | 0.01 |
| **Females** | **Non-carriers\*** | 144 | 24 (16.7) | 1.00 |  |  |
|  | **C282Y homozygotes** | 138 | 23 (16.7) | 0.99 | 0.53-1.87 | 1.00 |

\*Controls with no *HFE* C282Y or H63D alleles, matched on age, sex and BMI category.
Abbreviations: OR, Odds Ratio. CIs, Confidence Intervals.

### Sensitivity Analysis

#### Haemochromatosis

Excluding participants diagnosed with haemochromatosis at the time of iDXA strengthened the association between male C282Y homozygosity and vertebral fracture (OR: 3.37, 95% CI: 1.41-8.10, p=0.007), while estimates for females were unchanged (OR: 0.99, 95% CI: 0.53-1.90, p=1.00) (Table 4).

**Table 4.** Odds of vertebral fracture in *HFE* C282Y homozygotes compared to controls, excluding those with diagnosed haemochromatosis*.

| Group | *HFE* genotype | N | Vertebral Fracture, n (%) | OR | 95% CIs | p-value |
| --- | --- | --- | --- | --- | --- | --- |
| **Males** | **Non-carriers\*** | 90 | 9 (10) | 1.00 |  |  |
|  | **C282Y homozygotes** | 66 | 18 (27.3) | 3.37 | 1.41-8.10 | 0.007 |
| **Females** | **Non-carriers\*** | 144 | 24 (16.7) | 1.00 |  |  |
|  | **C282Y homozygotes** | 126 | 21 (16.7) | 0.99 | 0.53-1.90 | 1.00 |

\*Controls with no *HFE* C282Y or H63D alleles, matched on age, sex and BMI category.
Abbreviations: OR, Odds Ratio. CIs, Confidence Intervals.

#### Osteoporosis and Bone Mineral Density

Osteoporosis was assessed using HES records to identify participants with a diagnosis recorded before DXA imaging. Within the cohort, only seven participants had a recorded diagnosis of osteoporosis at the time of imaging, most of them female (Table 2). No male C282Y homozygotes had an osteoporosis diagnosis at the time of DXA, and no homozygotes with a vertebral fracture had a hospital-recorded diagnosis of osteoporosis.

Femoral neck BMD T-scores were lower among participants with vertebral fracture than among those without fracture, irrespective of genotype. However, within the fracture cohort, there was little evidence of a difference in mean BMD between C282Y homozygotes and matched controls. Among men with vertebral fractures, mean femoral neck T-scores were -0.82 in C282Y homozygotes and -0.83 in controls (mean difference 0.01, 95% CI -0.67 to 0.69). Among women with vertebral fractures, mean T-scores were -1.30 in C282Y homozygotes and -1.49 in controls (mean difference 0.19, 95% CI -0.43 to 0.81).

Males within the whole cohort had mean femoral neck T-scores of 0.25 in C282Y homozygotes and 0.52 in controls (mean difference, -0.27; 95% CI, -0.82 to 0.28), whereas in women they were -0.71 and -0.76, respectively (mean difference, 0.05; 95% CI, -0.32 to 0.42).

Importantly, despite the increased odds of vertebral fracture observed in male C282Y homozygotes, mean femoral neck T-scores remained within the WHO normal range (T-score > -1) in both male homozygotes and controls with vertebral fracture. In contrast, women with vertebral fractures had mean T-scores within the osteopenic range, regardless of genotype (Table 5). These findings suggest that the excess vertebral fracture burden observed in male C282Y homozygotes is not readily explained by differences in femoral neck BMD.

**Table 5:** Association between C282Y homozygosity and Bone Mineral Density.

|  |  | N | Cohort Mean Femoral Neck BMD T-score (SD) | Difference in mean BMD T-score† [95% CI] | N (with a vertebral fracture) (%) | Mean Femoral Neck BMD T-score (SD) (with vertebral fracture) | Mean BMD T-score difference with vertebral fracture† [95% CI] |
| --- | --- | --- | --- | --- | --- | --- | --- |
| Male | Non-carriers* | 82 | 0.52 (1.76) | -0.27 [-0.82 to 0.28] | 8 (9.76) | -0.83 (0.56) | +0.01 [-0.67 to 0.69] |
|  | C282Y Homozygotes | 80 | 0.25 (1.78) |  | 19 (23.75) | -0.82 (1.23) |  |
| Female | Non-carriers* | 134 | -0.76 (1.46) | +0.05 [-0.32 to 0.42] | 22 (16.42) | -1.49 (0.77) | +0.19 [-0.43 to 0.81] |
|  | C282Y Homozygotes | 130 | -0.71 (1.58) |  | 21 (16.15) | -1.30 (1.23) |  |

Abbreviations: BMD, Bone mineral density.
Differences in sample size across analyses reflect exclusions due to incomplete BMD data availability.
\* Non-carriers = no *HFE* C282Y or H63D alleles.
† Calculated as C282Y homozygotes minus non-carriers.

### Osteoarthritis and Back Pain

Among men without vertebral fracture, osteoarthritis was recorded in 8.6% of controls and fewer than 7.5% of C282Y homozygotes. In women without vertebral fracture, corresponding prevalences were 10.8% and 8.7%, respectively. Numbers were small among participants with vertebral fractures, preventing meaningful comparison by genotype.

Self-reported back pain lasting ≥3 months was more common among participants with vertebral fractures than among those without vertebral fractures. Among C282Y homozygotes, back pain was reported by <22.7% of men and <21.7% of women with vertebral fractures, compared with 14.9% and 15.7%, respectively, in those without vertebral fractures. However, there was no clear evidence of an association between C282Y genotype and back pain.

## Discussion

In this community-based cohort, male *HFE* C282Y homozygotes had almost threefold higher odds of vertebral fracture than matched controls without haemochromatosis-associated *HFE* alleles (OR 2.95, 95% CI 1.28–6.85), based on radiographically identified fractures on iDXA-based vertebral fracture assessment. By contrast, no excess fracture risk was observed among female homozygotes. Notably, many affected men had no prior diagnosis of haemochromatosis and mean femoral neck BMD remained within the WHO normal range, suggesting a burden of clinically unrecognised vertebral fractures that may not be identified through conventional BMD-based assessment.

Previous studies have demonstrated associations between haemochromatosis and reduced bone mass, osteoporosis and fracture risk, particularly in clinically diagnosed populations (33). However, genotype-specific evidence regarding vertebral fractures remains limited and inconsistent. A case-control study of 306 patients with haemochromatosis reported a non-significant increase in vertebral fractures overall, although fracture risk was higher in those with ferritin concentrations exceeding 1000 µg/L (25). Another cross-sectional study of 93 patients with haemochromatosis reported skeletal fragility in 20.4% of participants and noted a predominance of radiographic complications among C282Y homozygotes but did not specifically evaluate vertebral fractures by genotype (16). More recently, population-based studies have demonstrated increased fracture risk associated with both iron overload and *HFE* C282Y homozygosity, although vertebral fractures were identified through routine clinical records or studies focused on non-vertebral fractures (33,34). Our findings extend this evidence by demonstrating an increased prevalence of radiographically confirmed vertebral fractures in a genetically defined community cohort.

Comparable femoral neck BMD in male C282Y homozygotes and wild types contrasted with a significantly higher vertebral fracture risk observed in the homozygotes. Although limited by sample size, these findings suggest that factors other than areal BMD may contribute to skeletal fragility in this group. Iron overload and *HFE*-related dysregulation of the hepcidin-ferroportin axis are known to influence bone metabolism through increased osteoclast activity, inhibition of osteoblast function, oxidative stress, and chronic low-grade inflammation (27). Experimental and clinical studies suggest that these effects can compromise bone microarchitecture and material properties before measurable reductions in areal BMD occur (33). This microstructural deterioration may weaken vertebral bodies while DXA measurements remain within the normal range, offering a plausible explanation for the discordance between fracture risk and BMD observed in this study (36).

The association between C282Y homozygosity and vertebral fracture persisted after excluding individuals with a diagnosis of haemochromatosis. This suggests that increased fracture risk may not be confined to clinically recognised disease and could extend to individuals with undiagnosed or less severe iron overload. However, direct measures of iron burden and treatment history were unavailable, so these findings should be interpreted with caution.

The marked sex difference in our findings is consistent with the known lower penetrance and delayed clinical expression of haemochromatosis-related morbidity in women. While physiological blood loss may contribute to lower iron accumulation, hormonal regulation of iron metabolism is likely to play a central role. Oestrogen downregulates hepcidin expression and stabilises ferroportin activity, thereby limiting iron retention during premenopausal life and in women using hormone replacement therapy (37). Loss of this protective effect after menopause may partially explain the later onset of haemochromatosis-related complications in women and the absence of excess vertebral fracture risk observed in female homozygotes in this cohort.

Emerging evidence suggests that fracture risk in C282Y homozygotes may not be entirely attributable to tissue or cellular iron overload. Warny and colleagues followed 142,146 Danish general population individuals over a median of 11 years and observed markedly increased fracture risk in C282Y homozygotes with normal ferritin levels, including a higher cumulative incidence by age 80, which challenges the measurement of systemic iron as a method of triage for genotyping (34). Local iron deposition within bone, altered *HFE*-related cellular signalling and inflammatory pathways may contribute to skeletal fragility even in the absence of markedly elevated circulating iron markers (38). This may help explain why increased fracture risk has been reported in some C282Y homozygotes who do not meet clinical diagnostic criteria for haemochromatosis.

Vertebral fracture assessment using iDXA provides a practical, low-radiation method of identifying vertebral fractures during routine DXA examinations and is increasingly incorporated into fracture liaison services (39). Although conventional radiographs remain the reference standard, iDXA-based VFA has demonstrated good agreement with radiography for clinically relevant fractures (40,41). Although VFA may underestimate mild vertebral deformities compared with conventional radiography, all participants were assessed using the same imaging protocol. Any misclassification is therefore likely to be non-differential and expected to bias associations towards the null.

These findings have important clinical implications. Vertebral fractures are strong predictors of subsequent vertebral and non-vertebral fractures, (23), yet many remain undiagnosed. Our results suggest that men with C282Y homozygosity may represent a subgroup at increased risk of clinically silent vertebral fractures that is not identified through BMD assessment alone. Incorporating VFA into routine DXA assessment or fracture liaison services could therefore improve fracture detection and risk stratification in this population. The findings also support earlier identification of haemochromatosis and further investigation of whether treatment of iron overload can reduce skeletal complications, including occult vertebral fragility.

### Strengths and Limitations of the Study

The strengths of this study include the use of a large, well-characterised population cohort, genotype-defined exposure, blinded image review by an experienced reporter, and matched analysis to reduce confounding. The use of iDXA-based vertebral fracture assessment allowed identification of fractures not captured in routine clinical records, highlighting an otherwise hidden fracture burden. Several limitations should be acknowledged. UK Biobank participants are generally healthier than the wider population, which may limit the generalisability of the findings (42). Serum ferritin and transferrin saturation data were also unavailable, preventing direct assessment of iron loading.

## Conclusion

In conclusion, male *HFE* C282Y homozygotes in this community cohort had a higher prevalence of vertebral fractures detected by iDXA-based vertebral fracture assessment than those without *HFE* alleles, although BMD values appeared broadly similar between groups. These findings suggest that standard BMD-based assessment alone may underestimate vertebral fragility fracture risk in this group. The implications should be interpreted cautiously given the community-based UK Biobank sample, the absence of direct iron indices, and the observational design of this study. Our findings justify further evaluation of whether targeted vertebral fracture assessment and formal fracture-risk workup could improve detection and risk stratification in men with C282Y homozygosity.

## Data Availability

The data used in this study are available from UK Biobank to bona fide researchers upon application and approval. This research was conducted under UK Biobank application 14631. Restrictions apply to the availability of these data and they are not publicly available.

https://www.ukbiobank.ac.uk/

## Acknowledgements

This research has been conducted using the UK Biobank Resource, under application 14631. This work uses data provided by patients and collected by the NHS as part of their care and support, Copyright © (2023), NHS England. Re-used with the permission of the NHS England [and/or UK Biobank]. All rights reserved. The authors wish to thank the UK Biobank participants and coordinators for this unique dataset.

The authors would like to acknowledge the use of the University of Exeter High-Performance Computing (HPC) facility in carrying out this work.

For the purpose of open access, the author has applied a ‘Creative Commons Attribution (CC BY)’ licence to any Author Accepted Manuscript version arising from this submission.

## Authors contributions

LB: Conceptualisation; Data curation; Formal analysis; Methodology; Visualisation; Writing – original draft. JA: Conceptualisation; Data curation; Formal analysis; Writing – review & editing. LP: Data curation; Formal analysis; Writing – review & editing. KK: Conceptualisation; Writing – review & editing. DM: Conceptualisation; Writing – review & editing. JS: Writing – review & editing. LB, LP, and JA had access to the underlying data. All authors meet the ICMJE criteria for authorship, contributed to the interpretation of the results, and approved the final version of the manuscript.

